# Relative COVID-19 viral persistence and antibody kinetics

**DOI:** 10.1101/2020.07.01.20143917

**Authors:** Chung-Guei Huang, Ching-Tai Huang, Avijit Dutta, Pi-Yueh Chang, Mei-Jen Hsiao, Yu-Chia Hsieh, Shin-Ru Shih, Kuo-Chien Tsao, Cheng-Ta Yang

## Abstract

**Importance:** The COVID-19 antibody response is a critical indicator for evaluating immunity and also serves as the knowledge base for vaccine development. The picture is still not clear because of many limitations including testing tools, time of sampling, and the unclear impact of varying clinical status. In addition to these problems, antibody levels may not be equivalent to protective capacity.

**Objective:** To define the key factor for the different patterns of COVID-19 antibody response.

**Design:** We elucidated the antibody response with time-series throat and serum samples for viral loads and antibody levels, then used a neutralization test to evaluate protectiveness.

**Setting:** A medical center that typically cares for patients with moderate to severe diseases. Because of the low prevalence of COVID-19 in Taiwan and local government policy, however, we also admit COVID-19 patients with mild disease or even those without symptoms for inpatient care.

**Participants:** RT-PCR-confirmed COVID-19 patients.

**Results:** We found that only patients with relative persistence of virus at pharynx displayed strong antibody responses that were proportional to the pharyngeal viral load. They also had proportional neutralization titers per unit of serum. Although antibody levels decreased around 2 weeks after symptom onset, the neutralization efficacy per unit antibody remained steady and even continued to increase over time. The antibody response in patients with rapid virus clearance was weak, but the neutralization efficacy per unit antibody in these patients was comparable to those with persistent presence of virus. The deceased were with higher viral load, higher level of antibody, and higher neutralization titers in the serum, but the neutralization capacity per unit antibody is relatively low.

**Conclusions and Relevance:** Strong antibody response depends on the relative persistence of the virus, instead of the absolute virus amount. The antibody response is still weak if large amount of virus is cleared quickly. The neutralization efficacy per unit antibody is comparable between high and low antibody patterns. Strong antibody response contains more inefficient and maybe even harmful antibodies. Low antibody response is also equipped with a capable B cell pool of efficient antibodies, which may expand with next virus encounter and confer protection.

**Key points:** *Question:* The key factor for the different “patterns” of COVID-19 antibody response.

*Findings:* Strong antibody response depends on the relative persistence of the virus, instead of the absolute virus amount. The antibody response is still weak if large amount of virus is cleared quickly. The neutralization efficacy per unit antibody is comparable between high and low antibody patterns. High antibody level contains more inefficient antibodies.

*Meaning:* Strong response contains inefficient and maybe harmful antibodies. Low antibody response is also equipped with a capable B cell pool of efficient antibodies, which may expand with next virus encounter and confer protection.

## Main Text

In late January 2020, we started to treat RT-PCR (real-time polymerase chain reaction)-confirmed COVID-19 patients. We are a medical center that typically cares for patients with moderate to severe diseases. Because of the low prevalence of COVID-19 in Taiwan and local government policy, however, we also admit COVID-19 patients with mild disease or even those without symptoms for inpatient care. Serial RT-PCR tracking of pharyngeal samples was performed throughout each patient’s hospital course. With informed consent from patients or their families, our research was conducted using serum samples remaining after routine medical tests. We used two ELISA-based kits^1,2,3^ to detect anti-spike protein IgG antibodies, and the results of the two were concordant. The tests are semi-quantitative and measure antibody concentrations relative to a cut-off point value in serial dilutions of serum samples. We also cultured virus strains from our patient samples and used one of the strains to quantify the neutralization valence of serum samples in our Biosafety Level-3 laboratory. By mid-March 2020, we had collected serum samples from 15 consecutive patients. As a single center study, we also have complete medical record including detailed travel, occupation, contacts, and cluster history.

Theoretically, the stronger the viral stimulation, the stronger the immune response. We examined the viral load in the first pharyngeal specimens of the 15 patients, represented by E gene RT-PCR Ct value, and checked the association between the viral load and the highest antibody value in their serial serum samples. Seven patients had an absolute concordance. The higher the viral load, the stronger their antibody response, falling on a regression line with a correlation coefficient of 0.95 (Fig. 1). However, eight patients had no such correspondence. There was a clear difference in dynamic changes in the amount of virus at throat between these two groups (Fig. 2A). The virus persisted longer in the 7 patients with direct correlation, compared to the 8 patients with no correlation (21.3 ± 6.5 versus 12.0 ± 2.6 days, *p*=.015). The mean age of these two groups was also significantly different (60.1 ±10.3 years, median 65, versus 38.3 ± 14.9 years, median 38, *p*=.005).

**Figure 1.**
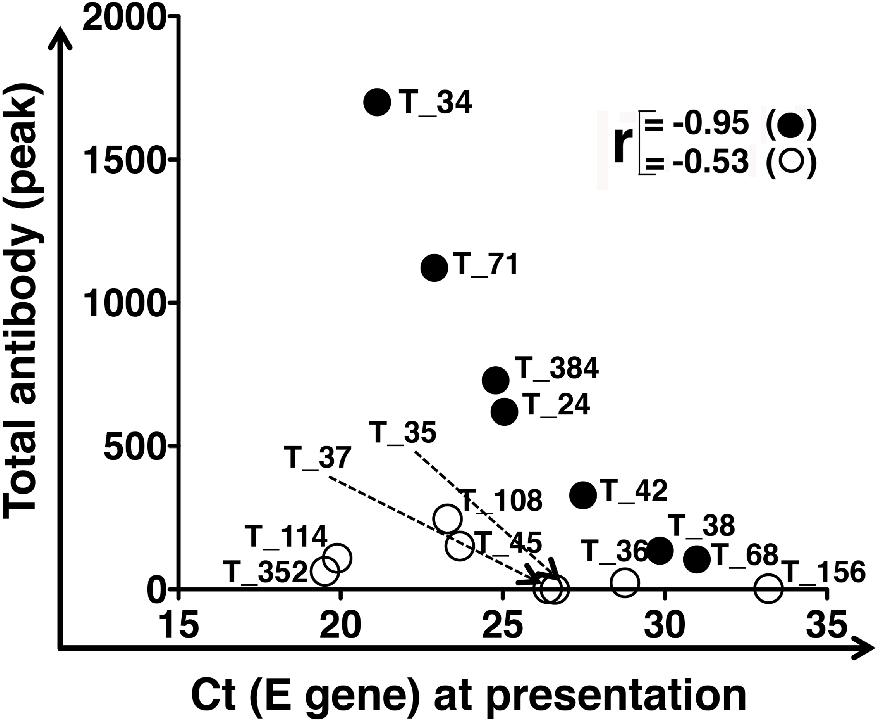
Maximal antibody response and pharyngeal virus load on disease presentation. The highest antibody levels were plotted in the Y-axis against the pharyngeal virus loads on disease presentation, as represented by the Ct values of viral E gene, in the X-axis. Seven patients in closed circles falls on a regression line of correlation coefficient 0.95. There is no correlation between the maximal antibody response and pharyngeal virus loads on disease presentation for the other 8 patients in open circles. The patients are tagged by our national serial number of COVID-19 cases.

**Figure 2.**
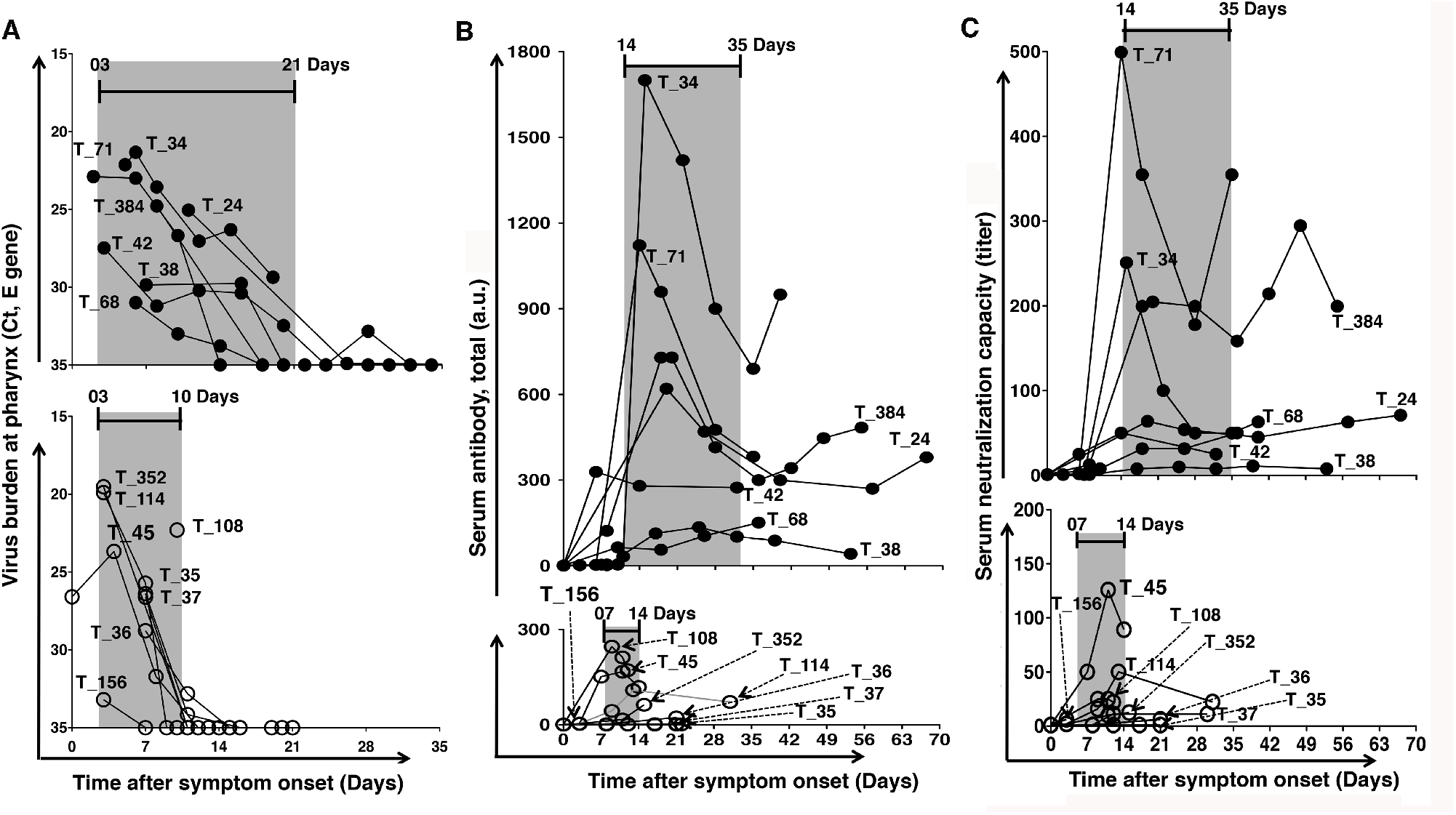
Dynamic changes in pharyngeal virus load, serum antibody and neutralization titer. **(A)** Seven patients with maximal antibody levels corresponding to virus loads on presentation (closed circles) revealed delayed clearance of virus from the throat. On the contrary, the other 8 cases with no correlation (open circles) eradicated the virus from the throat quickly. **(B)** The antibody response was significantly stronger in patients with delayed clearance compared to patients with rapid eradication. **(C)** The neutralization capacity was proportional to the amount of antibody in unit of serum. The neutralization valence was significantly higher in patients with persistent presence of the virus than patients with shorter presence of the virus. The patients are tagged by our national serial number of COVID-19 cases.

There were many more antibodies in patients with relatively persistent presence of virus than in those with rapid virus clearance (Fig. 2B). The highest antibody levels were 697.9 ± 384.2 and 74.8 ± 88.8, respectively (*p*=.026). The neutralization titer in each group was proportional to the amount of antibody (Fig. 2C). The peak neutralization titer was higher in patients with longer persistence of virus (162.5 ± 184.6 versus 29.7 ± 41.9, *p*=.090). High antibody levels were maintained for longer in patients with relative viral persistence. Antibody levels started decreasing 22.7 ± 8.6 days after symptom onset in this group, compared to 12.1 ± 5.6 days in those with rapid viral clearance (*p*=.019). The neutralization titers decreased accordingly.

We normalized the neutralization titer by the amount of antibody to determine the neutralization efficacy per unit of antibody. The neutralization efficacy per unit of antibody remained relatively steady or even continued increasing after two weeks (Fig. 3A), although the total antibody quantity and neutralization titer began to decline by that time. Interestingly, the neutralization efficacy per unit antibody was comparable between both groups, irrespective of viral persistence and antibody levels (41.3 ± 30.8 versus 35.7 ± 24.8, *p*=.704).

**Figure 3.**
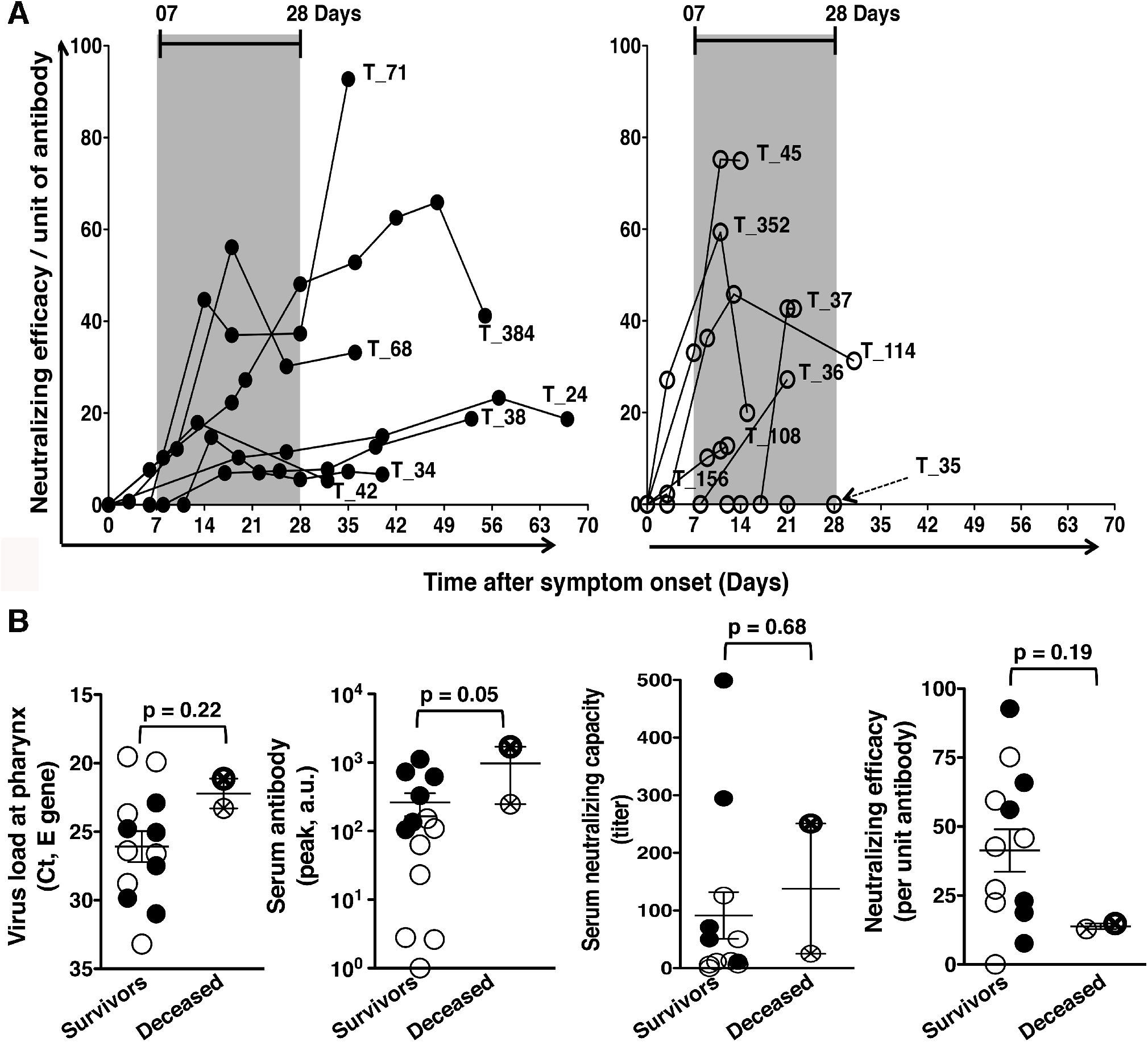
Neutralization efficacy per unit of antibody. **(A)** Even though the total antibody and neutralization capacity per unit of serum are significantly higher in patients with viral persistence, the neutralization efficacy per unit of antibody are comparable between patients with viral persistence (Left panel, closed circles) and patients with rapid eradication (Right panel, open circles). **(B)** The deceased (the two circles with a cross) were with high viral loads on presentation, higher amount and higher neutralization valence in unit of serum, but they had lower neutralization efficacy per unit of antibody compared to the survived (non-crossed circles, either closed or open).

Two out of our fifteen patients died. One young man was suspected to have suffered a thrombosis-related acute cardiac event. The other patient was older and had severe heart failure before contracting COVID-19. Although death is not necessarily a result of an inadequate immune response, the patients who died had a high amount of virus in the throat and higher level of antibodies in serum compared to those who survived, but the neutralization efficacy per unit antibody was relatively poor (Fig. 3B).

## Discussions

Many factors may contribute to viral persistence in the pharynx, including the size of virus inoculum and the immunity of the host. Slower clearance with relatively persistent presence of the virus induces strong antibody responses. Overall neutralization titers are associated with the amount of antibody per unit of serum. However, even though antibody levels begin to decline two week after symptom onset, the neutralization efficacy per unit of antibody remains the same or continues to increase. This indicates that the proportion of antibodies with lower neutralization efficacy gradually decreases, while the proportion of higher efficacy gradually increases. The phenomenon of neutralization efficacy increasing over time is in line with the known maturation process of the antibody response. Created through random VDJ recombination, the B cell receptor (BCR) repertoire is highly heterogeneous. Clonal selection is achieved through stimulation and response where B cells with BCR and antibodies with effective neutralization ability gradually expand and become the majority of the B cell pool responding to the virus. This is an evolutionary process that takes time. Neutralization capacity represents the antibody’s ability to protect against specific pathogens. It deserves special attention because there is a population of antibodies with poor neutralization capacity in the early stages of the antibody reaction. One of the most concerning risks of convalescent plasma therapy for COVID-19 is that some plasma antibodies may in fact not be protective and could even be harmful due to mechanisms such as Antibody-Dependent Enhancement (ADE).^4^ Therefore we must be cautious about the timing of plasma procurement from the patients who have recovered from the illness.

People tend to try to link the association between the amount of virus in respiratory samples to the severity of illness. However, persistent presence of virus rather than the absolute amount of virus at the throat was responsible for a strong and early antibody response. A strong and early antibody response is likely predominantly comprised of less protective and potentially even harmful antibodies. In patients with SARS, it has been reported that poor clinical outcomes was associated with early appearance of antibody^5^. Patients with difficulty eradicating the virus suffer from the damage caused by both the virus and the ineffective potentially harmful antibodies. In our study, patients with viral persistence and an earlier and stronger antibody response tend to be older. This may explain the vulnerability to COVID-19 in the elderly.

In our observations, low or even no detectable antibodies did not necessarily represent absence of immunity. Although the absolute antibody quantity in these patients is low, the neutralization efficacy per unit of antibody is equivalent to that of the group with higher antibody levels, indicating that patients with low antibody quantities also have a considerable number of mature B cells secreting effective antibodies. Upon subsequent encounters of the virus, these B cells will likely expand with a memory response and may produce effective antibodies in quantities sufficient to protect the host.

Among our patients, viral load at the throat did not persist for long in most healthcare workers (HCW). Their antibody response was also low. There are many possible reasons for this. These HCW are young. Personal protective equipment (PPE) may have reduced the amount of viral exposure. However, there is still significant morbidity and mortality from COVID-19 among HCW around the world. This may be due to insufficient personal protection or extremely high viral loads in their environment that overcome the protection afforded by PPE.

## Data Availability

All data are available in the manuscript. For any further details, correspondence should be addressed to Dr. Cheng-Ta Yang, Department of Thoracic Medicine, Chang Gung Memorial Hospital, Taoyuan, Taiwan. Phone: +886-3-3281200

## Methods

### CoVID-19 Nucleic acid detection

Nasopharyngeal or oropharyngeal throat swab specimens were collected from patients. Test for COVID-19 nucleic acid followed standard protocols. RNA was extracted from clinical samples with the LabTurbo system (Taigen, Taiwan). A 25 μL reaction contained 5 μL of RNA, 12.5 μL of 2 × reaction buffer provided with the Superscript III one step RT-PCR system with Platinum Taq Polymerase (AgPath-ID One-step RT-PCR Kit), 1 μL of reverse transcriptase/Taq mixture from the kit, 0.4 μL of a 50 mM magnesium sulphate solution (Invitrogen), and 1 μg of nonacetylated bovine serum albumin (Roche). All oligonucleotides were synthesized and provided by Tib-Molbiol (Berlin, Germany). Thermal cycling was performed at 48 °C for 30 min for reverse transcription, followed by 95 °C for 10 min and then 45 cycles of 95 °C for 10s, 65 °C for 30s.

## COVID-19 Serum antibody detection

To evaluate the antibody response, the levels of total IgG in patients’ sera were semi-quantified by ELISA (cat No. WS-1096, WANTAI SARS-CoV-2 Ab ELISA, China). WANTAI SARS-CoV-2 Ab ELISA is a two step incubation antigen “sandwich” enzyme immunoassay kit, which uses polystyrene microwell strips pre-coated with recombinant SARS-CoV-2 antigen. Patient’s serum or plasma specimen is added, and during the first incubation, the specific SARS-CoV-2 antibodies will be captured inside the wells, if present. The microwells are then washed to remove unbound serum proteins. Second recombinant SARS-CoV-2 antigen conjugated to the enzyme Horseradish Peroxidase (HRP-Conjugate) is added, and during the second incubation, the conjugated antigen will bind to the captured antibody inside the wells. The microwells are then washed to remove unbound conjugate, and Chromogen solutions are added into the wells. In wells containing the antigen-antibody-antigen(HRP) “sandwich” immunocomplex, the colorless Chromogens are hydrolyzed by the bound HRP conjugate to a blue colored product. The blue color turns yellow after the reaction is stopped with sulfuric acid. The amount of color intensity can be measured and it is proportional to the amount of antibody captured inside the wells, and to the specimen respectively. Wells containing specimens negative for SARS-CoV-2 antibodies remain colorless. The results had been verified with another ELISA kit (Anti-SARS-CoV-2 ELISA IgG, Euroimmun, Germany), the ELISA results got good agreement in both kits.

## Neutralization Antibody Test (NAT)

The neutralizing antibody test of COVID-19 followed the standard protocol of a plaque reduction neutralization test. Vero cells were regularly maintained in minimal essential medium (MEM) supplemented with 10% (v/v) fetal bovine serum (FBS). COVID-19 virus was propagated in Vero cells in maintenance medium consisting of MEM supplemented with 0% FBS. Serum samples were inactivated at 56°C for 30 min prior to use. Serial two-fold dilutions of sera were mixed with an equal volume of COVID-19 virus suspension containing 100 × the median tissue culture infectious dose (TCID_50_). The mixture was incubated for 2 hr at 37°C and then an equal volume of suspended VeroE6 cells (approximately 30,000 cells/well) were added to each well. Following incubation for 1 week at 37°C, cells were fixed with 5% glutaraldehyde and stained with 0.1% crystal violet. Serum neutralization titers were calculated and expressed as the reciprocals of the highest serum dilution that inhibits cytopathic effects.

## Statistical analysis

We used Graph Pad Prism version 5 for statistical analyses. Data represented as mean ± SD and *p* values for two-tailed unpaired Student’s t-test in the text.

